# Survival trends among people living with human immunodeficiency virus on antiretroviral treatment in two rural districts in Ghana

**DOI:** 10.1101/2023.08.17.23294224

**Authors:** Eugene Sackeya, Martin Muonibe Beru, Richard Nomo Angmortey, Kingsley Boakye, Musah Baatira, Mohammed Sheriff Yakubu, Douglas Aninng Opoku, Aliyu Mohammed, Nana Kwame Ayisi-Boateng, Daniel Boateng, Emmanuel Kweku Nakua, Anthony Kweku Edusei

## Abstract

**Background:** The human immunodeficiency virus (HIV) has caused a lot of havoc since the early 1970s, affecting 37.6 million people worldwide. The 90-90-90 treatment policy was adopted in Ghana in 2015 with the overall aim to end new infections by 2030, and to improve the life expectancy of HIV seropositive individuals. With the scale-up of Highly Active Antiretroviral Therapy (HAART), the lifespan of People Living with HIV (PLWH) on antiretrovirals (ARVs) is expected to improve. In rural districts in Ghana, little is known about the survival probabilities of PLWH on ARVs hence, this study was conducted to estimate the survival trends of PLWH on ARVs for the periods between 2016 to 2020.

**Methods:** A retrospective evaluation of data gathered across ARV centers within Tatale and Zabzugu districts in Ghana from 2016 to 2020 among PLWH on ARVs. The census technique was employed and a total of 261 participants were recruited for the study. The data was analyzed using STATA software version 16.0. Lifetable analysis and Kaplan-Meier graph were used to assess the survival probabilities. “Stptime” per 1000 person-years and the competing risk regression was used to evaluate mortality rates and risk of mortality.

**Results:** The cumulative survival probability was 0.8847 (95% CI: 0.8334-0.9209). The overall mortality rate was 51.89 (95% CI: 36.89-72.97) per 1000 person-years. WHO stage III and IV [AHR: 4.25 (95%CI: 1.6-9.71) p = 0.001] as well as age group (50^+^ years) [AHR: 5.02 (95% CI: 1.78-14.13) p=0.002] were associated with mortality.

**Conclusion:** Survival probabilities is high among the population of PLWH in Ghana with declining mortality rates. Clinicians should provide critical attention and care to patients at HIV WHO stages III and IV and intensify HIV screening at all entry points since early diagnosis is associated with high survival probabilities.

## Background

The human immunodeficiency virus (HIV), the organism responsible for acquired immune deficiency syndrome (AIDS) has caused a lot of havoc to individuals and families since its outbreak in the early 1970s [1,2]. HIV has affected 37.6 million people worldwide as of 2020, with an estimated 27.4 million people accessing antiretroviral (ARVs) therapy globally [3].

The majority affected by this HIV pandemic live in low and middle-income countries (LMICs) with 20.6 million (55%) of all cases from eastern and southern Africa, while 4.7 million (13%) are in western and central Africa [3,4]. In Ghana, about 342,307 people are living with HIV (PLWH) and of these, 77% of them are receiving lifesaving highly active antiretroviral therapy (HAART) [5]. The therapeutic value of antiretroviral therapy (ART) for PLWH is undeniable and early initiation of ART reduces morbidity and mortality. It is evident that when HIV replication is suppressed, patients have significant improvement in immunologic status, as seen by higher clusters of differentiation 4 (CD4) counts, lower AIDS-related morbidity and death, and, in many cases, a return to a normal or near-normal quality of life [6]. The advantages of early ART include a reduction in not just classic AIDS-related problems, but also end-stage organ damage as well as non-AIDS-defining malignancies [6].

ART administration was introduced in Ghana in June 2003 as part of a comprehensive care package including voluntary counseling and testing (VCT), prevention of mother-to-child transmission (PMTCT) of HIV, and the treatment and management of sexually transmitted infections (STIs) [7]. However, only PLWH whose CD4 count was ≤ 350 or classified as being in WHO stages III and IV were initiated on ARVs [7,8]. Owing to the global impact of the HIV pandemic exceeding all expectations, with a mortality peak of 1.9 million deaths in 2006 to 0.95 million in 2017 [9], the World Health Organization (WHO) and its partners adopted an ambitious treatment target, dubbed the 90-90-90 agenda which was reviewed in 2020 to 95-95-95. In this policy, 95% of PLWH should know their status, 95% of those who know their status should be on ARVs and 95% of those on ARVs should achieve viral suppression [4]. This treatment policy is in line with sustainable development goal three which emphasizes the need for good health and well-being of all people by 2030 with HIV being one of the significant indicators [11].

Ghana, a WHO member state adopted this policy in 2015 with efforts made to increase accessibility to HIV counseling and testing services, provision of ARVs for all those living with HIV and enhanced evaluation of effectiveness of ARVs through regular viral load assessment at regular intervals at no cost to the clients [5]. As of 2019 in Ghana, 58% of PLWH know their HIV status, of this number, 77% of them are receiving ARVs and 68% of those receiving ARVs have their viral load suppressed [5,12].

During the early 1980s, to be diagnosed with HIV in Ghana was synonymous with a death sentence, however, with the scale-up of HAART, more PLWH are now receiving the lifesaving ARVs with the aim of improving their life expectancy [13]. In Ghana there is limited data on survival trends of PLWH. This study therefore sought to estimate the survival trends of PLWH on ARVs for the periods between 2016 to 2020, the mortality rate and the associated risk factors.

## Methods

### Study Design

The study was a retrospective evaluation of data from antiretroviral therapy (ART) sites in two rural districts in Ghana (Zabzugu and Tatale Districts).

### Study Setting

The study was conducted in the Zabzugu and Tatale districts of the northern region of the Republic of Ghana. These two districts were purposively selected based on their active involvement in ART in the region. Participants were drawn from four ART centers at the Zabzugu district hospital (ZDH), Tatale district hospital (TDH), Kpalbutabu Health Center -Tatale and Nakpali Health Center -Zabzugu. With the capitals of the two districts being Zabzugu and Tatale respectively, they share a border with Togo in the east, to the north with Saboba, to the west Yendi, and to the south, Wulensi. Zabzugu District covers an area of 1,100.1 square kilometer and Tatale Sangule district covers an area of about 1,130 square kilometers [14,15].

### Study Population

The study population included PLWH, 13 years old and above, in the Tatale and Zabzugu districts who were diagnosed and enrolled on ARVs. The study participants were PLWH who were diagnosed not earlier than 2016 or were diagnosed earlier but started ARVs in 2016 through to 2020. PLWH less than 13years at the time of registration due to their dependency on parents and care givers likely to influence adherance and those who were transferred from districts outside the two districts for lack of adequate information were excluded.

### Sampling Technique

The census sampling technique was used for the selection of the research participants. The study participants included all PLWH who were diagnosed with HIV from 2016 to 2020 and followed for varying number of years, but not exceeding five years. A sample size of 261 participants were considered for this study. The sample size constituted the entire number of PLWH who started their treatment from 2016 to 2020. A total of 275 PLHIV were registered within the period, while 14 participants were excluded from the study as a result of transfers, under age (< 13years) and incomplete information. Most of the participants entered the study at different entry points but 31st December 2021 was the administrative censoring date for all participants.

### Variables description

#### Dependent variable

The main outcome variable was the survival probabilities of PLWH on ARVs. This predicted the chance of a participant being alive following the initiation to ARVs during the study period. The secondary outcomes included the mortality rate and the risk factors for mortality.

#### Independent variables

The study used eleven (11) independent variables. These included the type of regimen the person was on, which is either a efavirenz-based combination or a dolutegravir-based combination, comorbidity/coinfection defined as any other medical condition that co-existed with HIV or AIDS during diagnosis, HIV status disclosure refers to disclosing one’s HIV status to anyone other than a healthcare provider, and enrolment on the National Health Insurance Scheme (NHIS). The remaining variables included the participant’s marital status, WHO HIV stages (I, II, III, and IV), client’s locality or area of residence, client’s health facility where ARVs is received, gender and religion.

### Data Collection and Management

Data collection was conducted from 1^st^ July 2022 to 31^st^ August 2022 using a standard checklist by two public health nurses and a postgraduate student of Epidemiology and Biostatistics. This was done by reviewing existing medical records including the patient’s ARVs folder, the pharmacy logbook, the ARVs logbook, and the viral load register. Two different data collection officers collected the same data set and incomplete forms were returned to be completed and those that were not completed for lack of adequate information were dropped. The data was validated by comparing the data from the two research assistants to ensure accuracy and consistency. After cleaning, the data was stored in a hard drive and also in Google drive for future use and protection.

### Data Analysis

Data was analyzed using the STATA statistical software version 16.0. Descriptive statistics were used to describe the baseline characteristics of participants including the socio-demographic and the clinical characteristics. For the person-time computation, the date of administrative censorship (December 31, 2021) was considered as the last date for all persons who were still alive and receiving treatment. For the assessment of the survival probabilities, lifetable analysis was done alongside the display of a survival function using a Kaplan-Meier graph to assess variations in survival among subgroups. The overall mortality rate was calculated using stptime per 1000 person years. The stptime command computed and tabulated the person-time and the incidence rate. The subgroup mortality rates were calculated using stptime adjusting for the subgroup per 1000 person years. To examine the association between baseline factors and mortality, the Competing risk regression model was used. Statistical significance was set at p-value <0.05 at a confidence interval of 95%. Variables (both significant and insignificant at the Univariate level) that were deemed to be important based on previous studies [15,16] were included in the multivariable analysis.

### Ethical consideration

Ethical clearance was granted by the Committee for Human Research, Publication and Ethics of the Kwame Nkrumah University of Science and Technology with reference number CHRPE/AP/287/22. Participants informed consent was not taken since this study involved a review of secondary data from hospital medical records and the patients were not available at the time of the data collection. This was explained in the application for the ethical clearance and participants’ consent was waived by the ethics committee. All the study data that were collected were completely anonymized.

## Results

### Socio-Demographic Characteristics of Participants

The study participants who were in the age category of 30 to 39 years were 41.0% with the median age (interquartile range) of 35 (14) years. More than half (66.28%) of study participants were females while a little over one third (42.53%) were Christians. Approximately 51.34% resided in the Tatale Sanguli district even though 58.62% received their medications from the Zabzugu district. Those who had no formal education constituted 72.03% of the participants while 61.30% were non-skilled workers. Approximately 89.7% were married and 90.42% of them were registered with the National Health Insurance Scheme (NHIS) (Table 1).

**Table 1:** Socio-demographic characteristics of participants.

| **Variable** | **Zabzugu District**<br>**Frequency (%)** | **Tatale District**<br>**Frequency (%)** | **Total (%)** |
| --- | --- | --- | --- |
| Age (years) |  |  |  |
| **13-19** | 2 (0.8) | 10 (3.8) | 12 (4.6) |
| **20-29** | 32 (12.3) | 31 (11.9) | 63 (24.2) |
| **30-39** | 74 (28.4) | 33 (12.6) | 107 (41.0) |
| **40-49** | 28 (10.7) | 14 (5.4) | 42 (16.1) |
| **50+** | 17 (6.5) | 20 (7.6) | 37 (14.1) |
| Median age (IQR) | **35 (14)** |  |  |
| Gender |  |  |  |
| **Male** | 59 (22.6) | 29 (11.1) | 88 (33.7) |
| **Female** | 94 (36.0) | 79 (30.3) | 173 (66.3) |
| Religion |  |  |  |
| **Christain** | 50 (19.2) | 61 (23.1) | 111 (42.5) |
| **Muslim** | 64 (24.5) | 17 (6.5) | 81 (31.1) |
| **Traditional** | 39 (14.9) | 30 (11.5) | 69 (26.4) |
| District-of-Residences |  |  |  |
| **Zabzugu district** | 118 (45.2) | 1 (0.4) | 119 (45.6) |
| **Tatale district** | 29 (11.1) | 105 (40.2) | 134 (51.3) |
| **Other districts** | 6 (2.3) | 2 (0.8) | 8 (3.1) |
| Educational Level |  |  |  |
| **Formal education** | 36 (13.8) | 37 (14.2) | 73 (28.0) |
| **No formal education** | 117 (44.8) | 71 (27.2) | 188 (72.0) |
| Occupation |  |  |  |
| **Unemployed** | 50 (19.2) | 30 (11.5) | 80 (30.7) |
| **Skilled worker** | 12 (4.6) | 9 (3.5) | 21 (8.1) |
| **None skilled worker** | 91 (34.9) | 69 (26.4) | 160 (61.3) |
| Marital Status |  |  |  |
| **Never Married** | 10 (3.8) | 17 (6.5) | 27 (10.3) |
| **Ever Married** | 143 (54.8) | 91 (34.9) | 234 (89.7) |
| NHIS Status |  |  |  |
| **Registered with NHIS** | 139 (53.3) | 97 (37.1) | 236 (90.4) |
| **Not registered with NHIS** | 14 (5.4) | 11 (4.2) | 25 (9.6) |

NHIS = National Health Insurance, ZDH = Zabzugu District Hospital, TDH = Tatale District Hospital, IQR: Interquartile range

### Clinical characteristics of the study participants

Table 2 presents results of clinical charcteristics of the study participants. Majority (64.0%) were diagnosed at WHO stage I and 42.91% had disclosed their HIV status to another person other than the healthcare provider. Most of the participants (83.14%) were on Efavirenz-based Combination while 52.87% were alive and traceable at the time of the study.

**Table 2:**
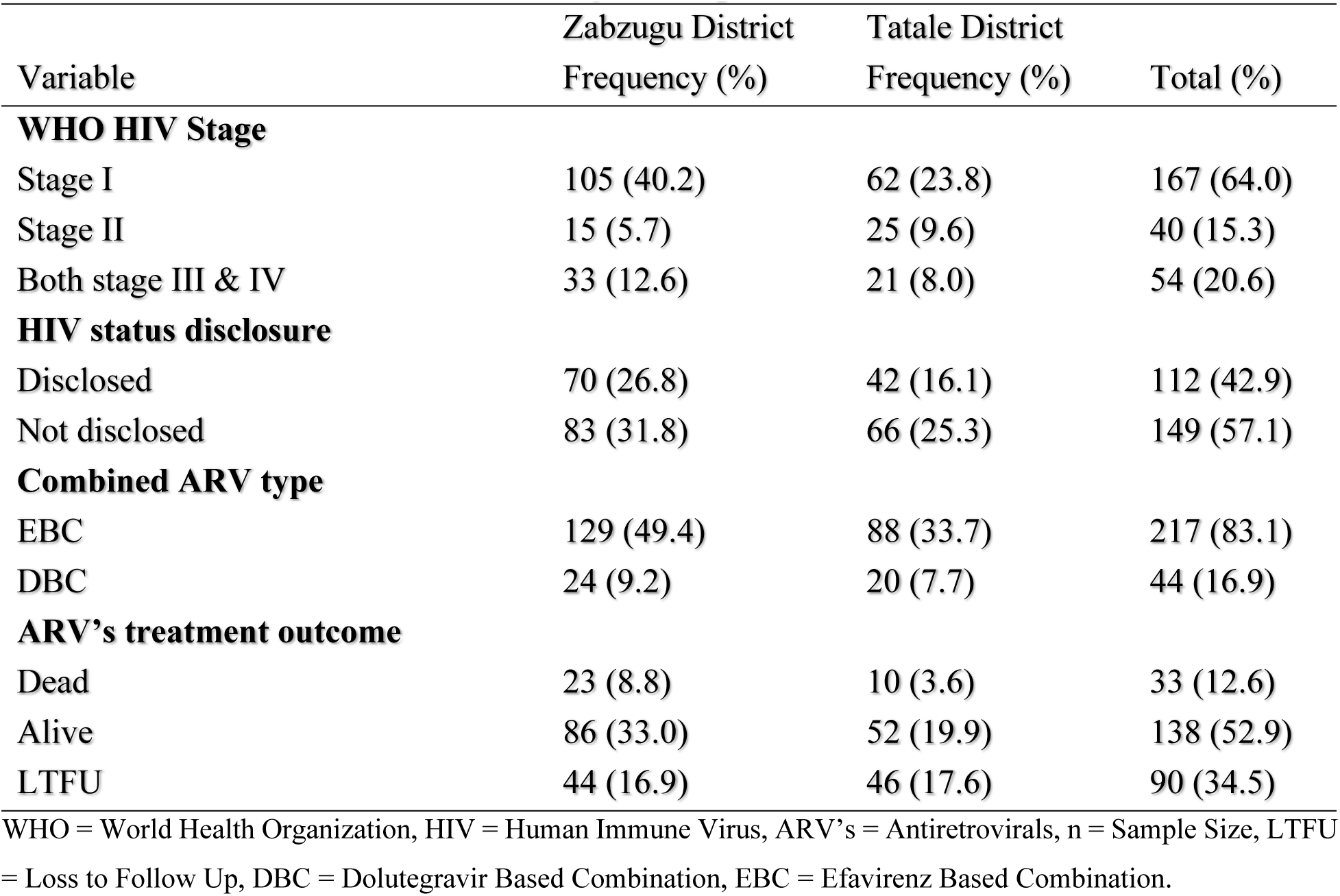
Clinical Characteristics of Study Participants.

### Five-Year Trend of Survival Probabilities

A total of 635.981 person-years with 2.17 median person years of follow-up was contributed by the 261 participants. In the first year of follow-up, 15 mortalities were recorded giving a survival probability of 0.9376 (95% Cl: 0.8987-0.9619) for the period. After three years of follow-up, the cumulative mortality was 32 and a survival probability of 0.8337 (95% CI: 0.7704-0.8809). With decline in mortalities over the years, the overall mortality at the end of the fifth years was 33 and a survival probability of 0.8049 (95% CI: 0.7145-0.8693) (table 3).

**Table 3:**
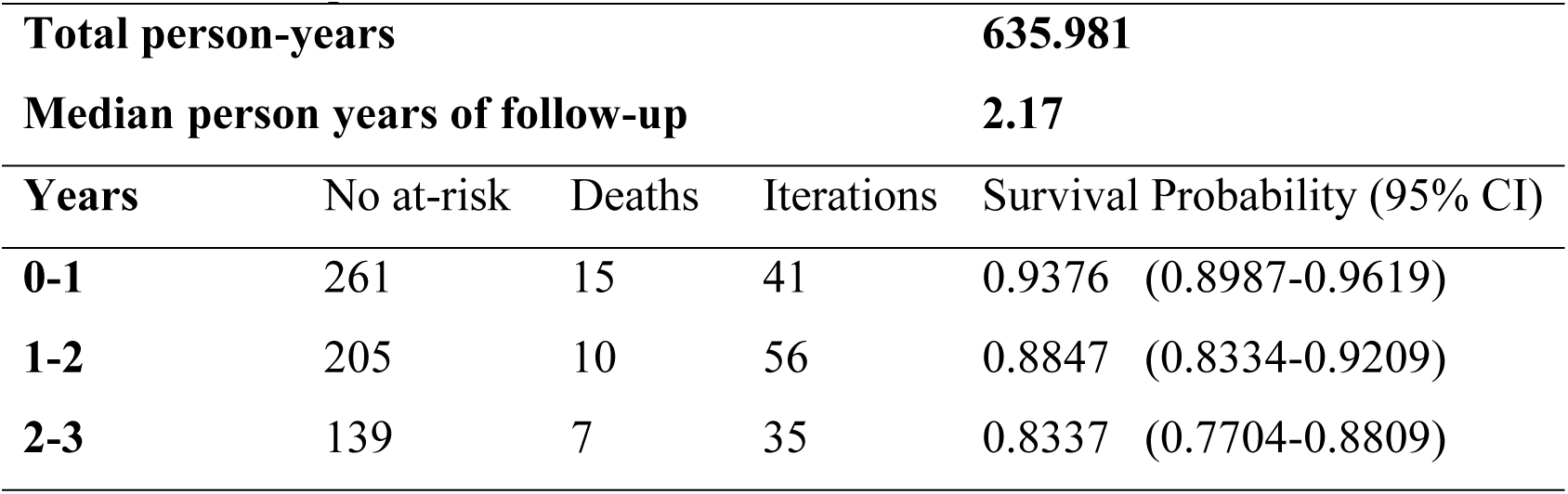
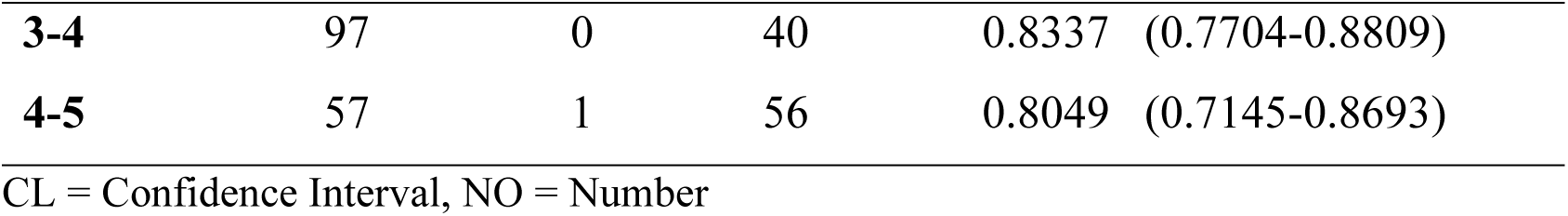
survival probabilities.

Overall, the Kaplan-Meier survival curve revealed that the majority of fatalities occurred in the first to third years after starting ARVs, thus 15, 10 & 7 respectively. Even though the overall cumulative survival probability for the five-year study period was 0.8049, those who were diagnosed at the WHO stages I and II had a higher survival probability as shown in figure 1 and so was for women and those in the age group of 13-29 years. Patients who did not disclose their HIV status had higher survival probability than those who disclosed their status but this trend changed after two years of follow-up where the survival probabilities of those who disclosed their status improved (Figure1).

**Figure 1:**
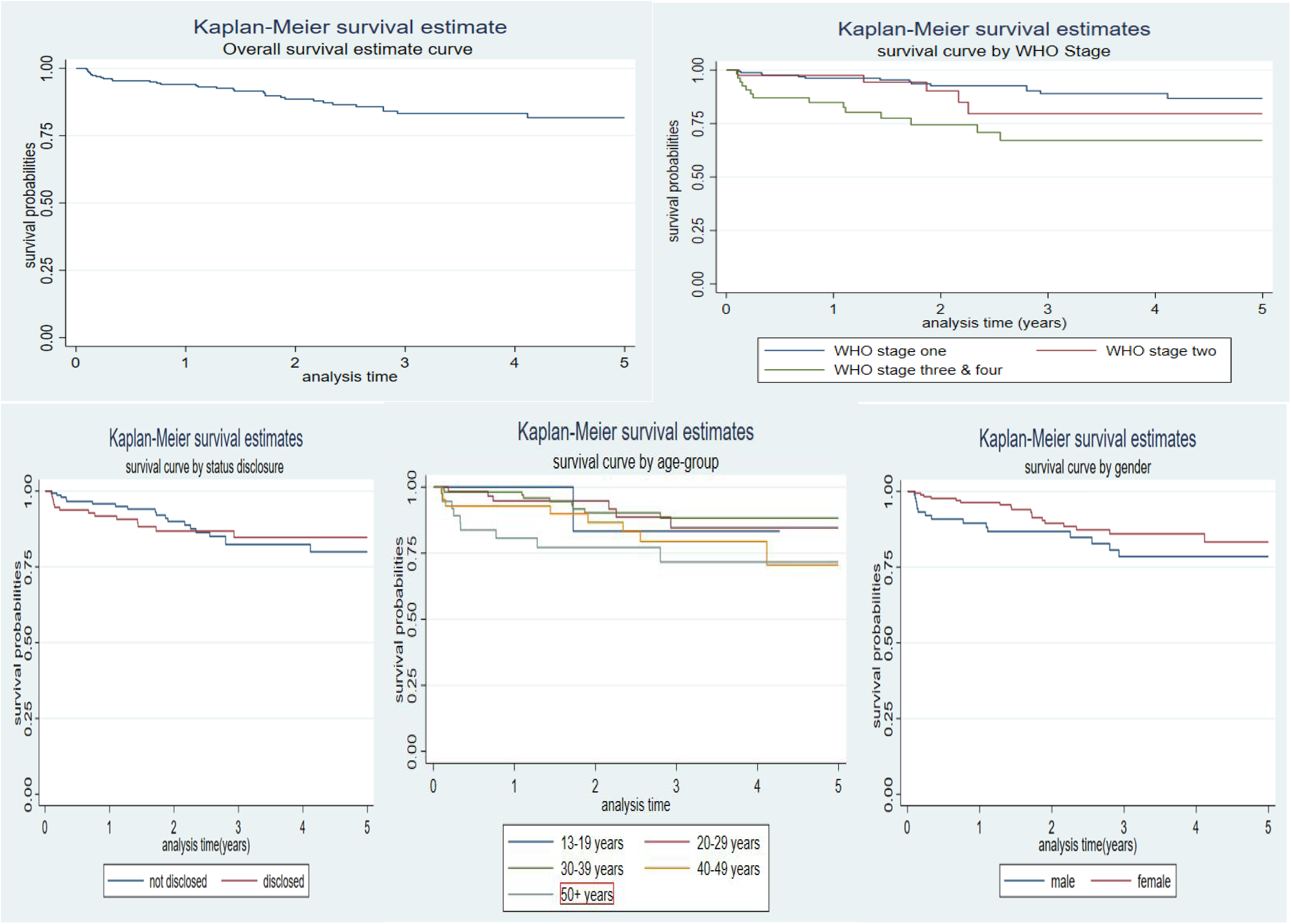
Kaplan Meier’s Survival Estimates

### Five-Year Mortality Rates among PLWH on ARVs

Thirty-three (12.64%) participants died during the study period. The total person years for the five-year period was 635.98 person years with the overall mortality rate of 51.89 (95% CI: 36.89-72.97) per 1000 person years. Mortality rates were higher above the overall rate among participants 30 years and above, 55.22 (95% CI: 37.60-81.10) per 1000 person years and males 68.65 (95% CI: 41.39-113.87) per 1000 person years. The mortality rate was almost even among those who disclosed their HIV status and those who did not with 53.60 (95% CI: 31.75- 90.51) per 1000 person-years and 50.69 (95% CI: 32.34-79.48) per 1000 person-years respectively. WHO stage III and IV recorded the highest mortality rate of 123.75 (95% CI: 73.29-208.95) per 1000 person-years.

In the univariate competing risk regression analysis, age was statistically significant and associated with mortality. The hazard of mortality was 3.28 times higher among those aged 50 and above [HR: 3.28; 95% CI: 1.29-8.33, p=0.013] and after adjusting for confounders in the multivariable analysis [AHR: 5.02; 95% CI: 1.78-14.13, p=0.002]. WHO stage was statistically significant, the hazard of mortality was 3.51 times higher among those diagnosed at WHO stage III & IV [HR: 3.51; 95% CI: 1.67-7.38, p=0.001] and the same was the case after adjusting for confounders [AHR: 4.63; 95% CI: 1.91-11.18, p=0.001], however, we cannot rule out region and marital status as factors influencing mortality among PLWH (Table 4).

**Table 4:**
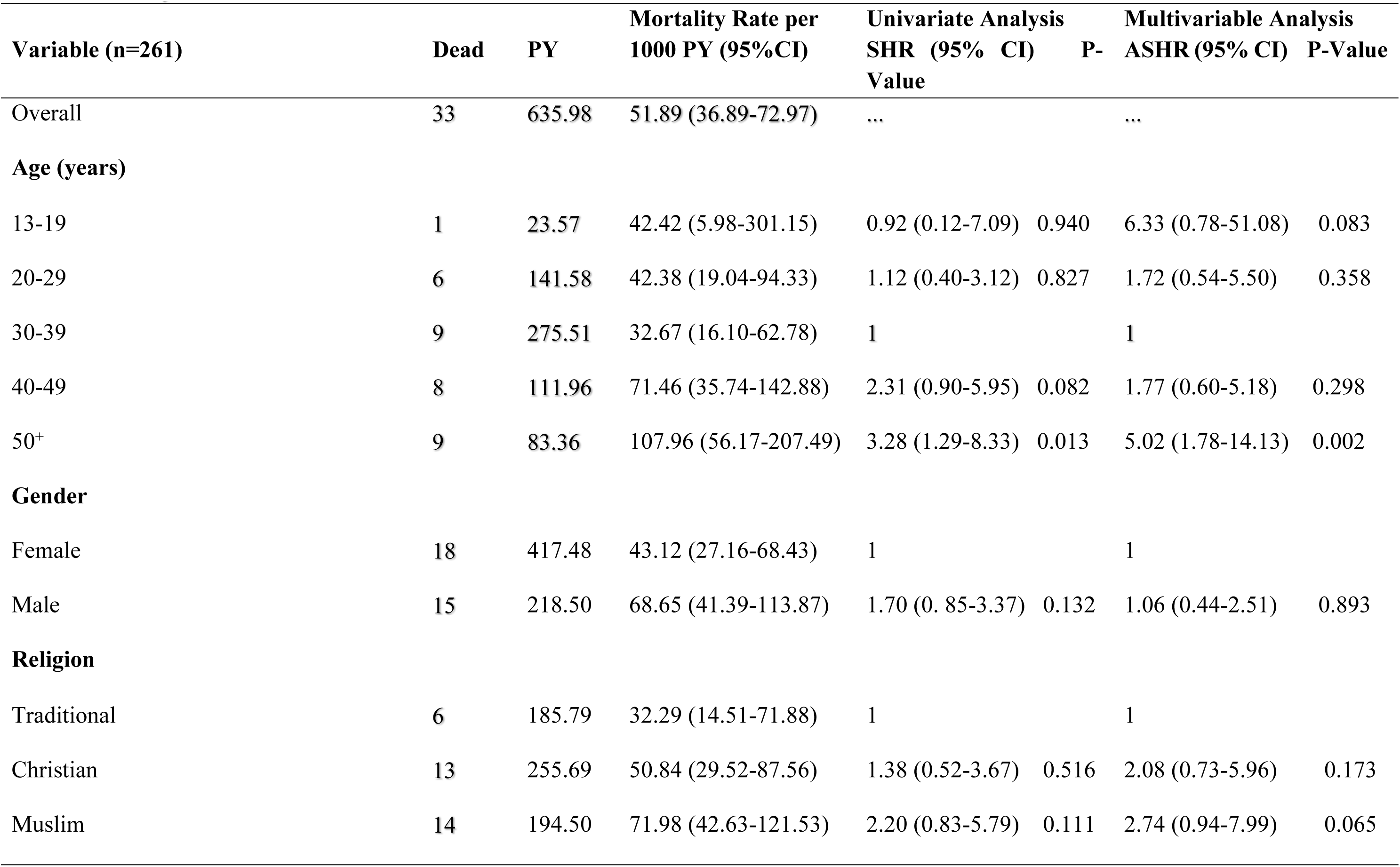
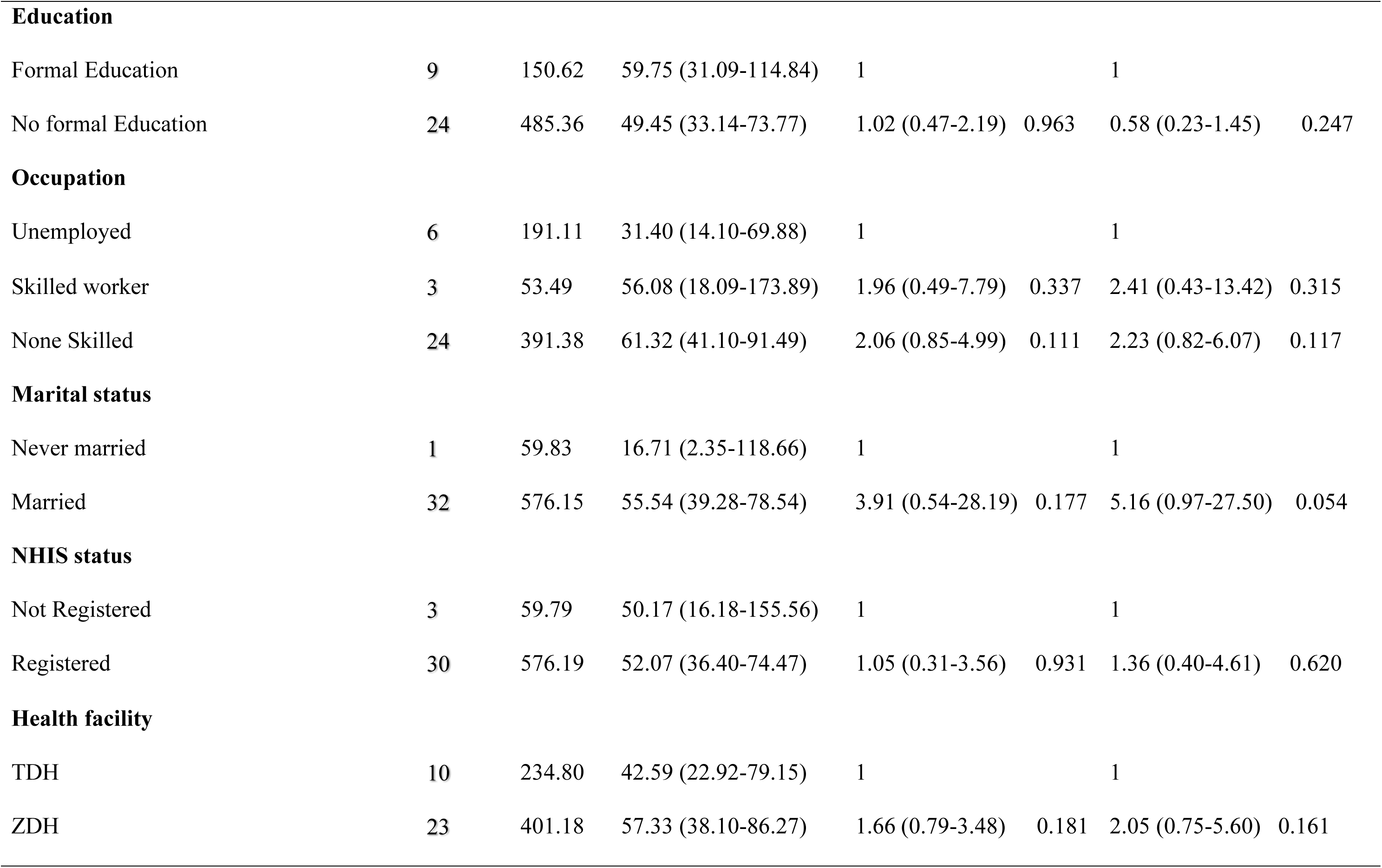
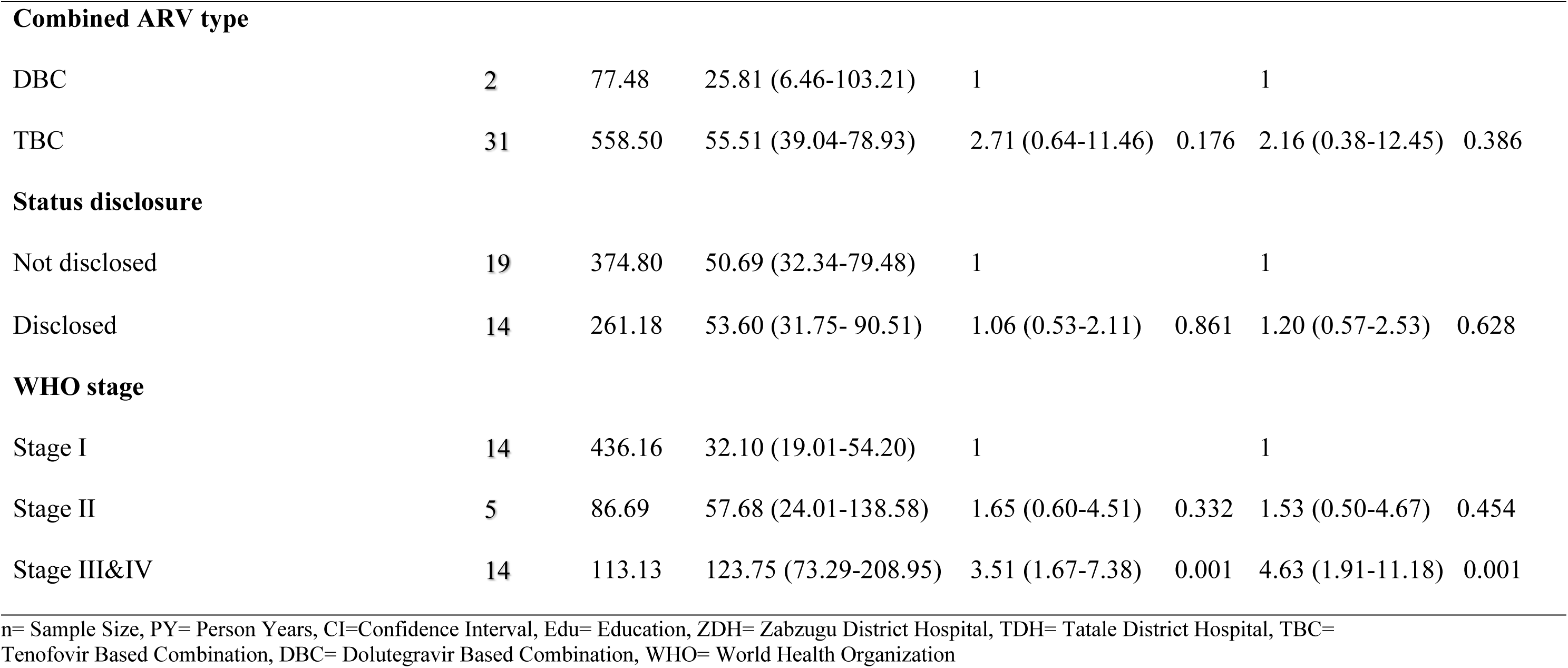
Mortality rates and their determinants.

Figure 2 presents the illustration of the hazard of mortality over the study period. The hazard of mortality is highest between the first and the second year of follow-up but declined steadily over the rest of the study period from the second year on wards.

**Figure 2:**
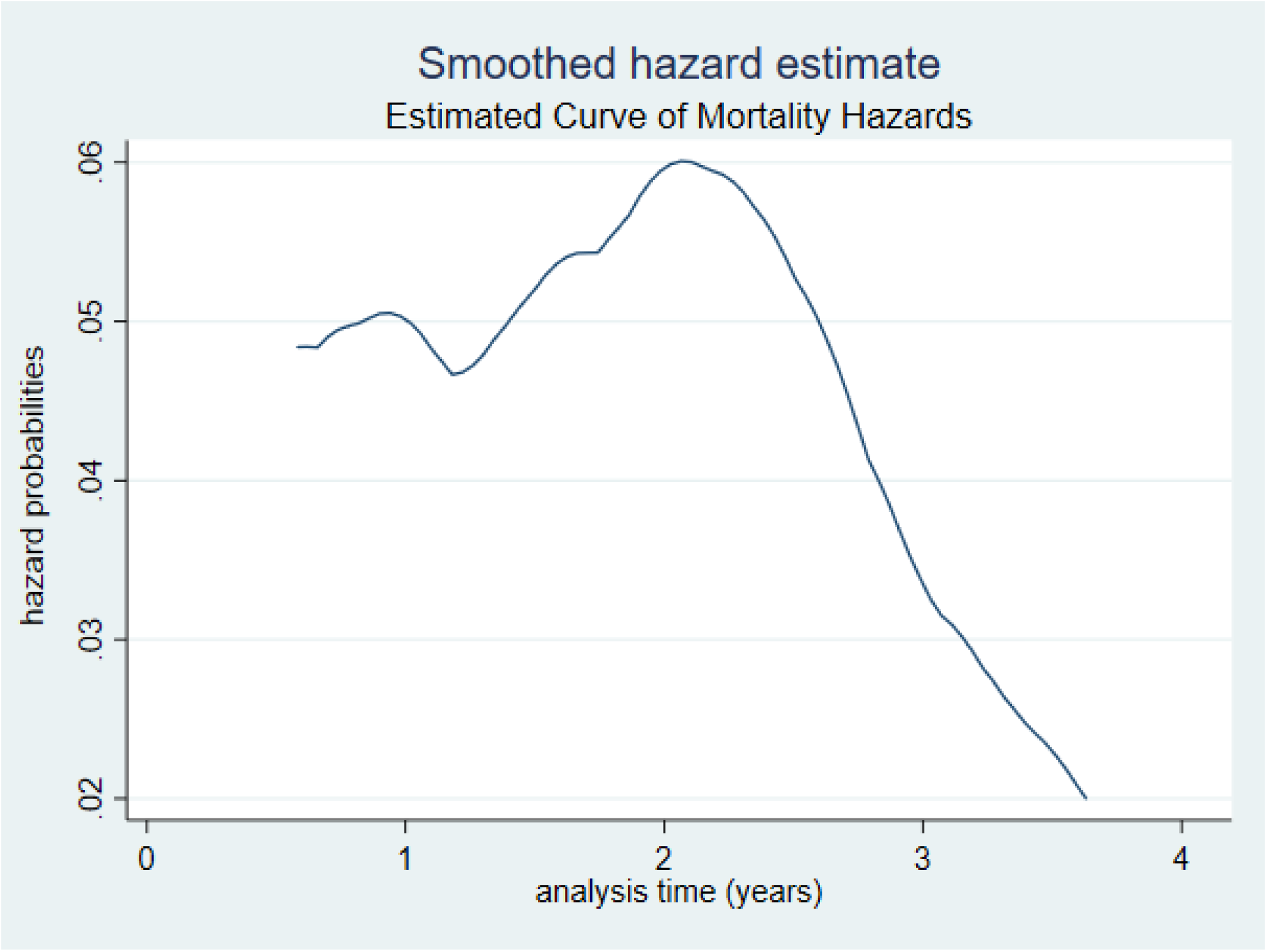
Mortality Hazard curve

## Discussion

Preventing major AIDS-related and non-AIDS-related illnesses in HIV-infected individuals requires early initiation of antiretroviral medication [16]. The majority of the participants were initiated to ARVs at WHO stages I and II (79.31%), This was however different from what was reported in a study conducted in Ghana to assess the effects of antiretroviral therapy on all-cause of mortality among PLWH. In that study, 51.0% of PLWH were diagnosed at WHO stage I and II [17]. An expanded study involving most of the HIV sentinel sites in Ghana also found an even lower (27.7%) prevalence of HIV initiation at WHO stage I & II [18]. The variation in findings could be due to the fact that the study was conducted during the period of the 90-90-90 treatment policy where there was a more accelerated action for HIV testing and treatment. During that period, testing was done at the OPD instead of the laboratory where only clients who were suspected of being infected were referred for testing. Providing testing services for all persons, whether symptomatic or not, meant that more clients would be diagnosed in the earlier stages of the disease.

The current study identified the majority were alive and on treatment at the end of the five years of follow-up, while 34.5% were lost to follow up (LTFU) and the remaining 12.64% were dead. Our findings were a little different from the findings in a study in a primary public hospital of Wukro, Tigray, Ethiopia, where 11% of LTFU was recorded [19]. The reason for the high rate of LTFU could be due to the high numbers diagnosed at WHO stages I and II, who are asymptomatic and may not see the need to take their medications. Most of the fatalities occurred in the first and second years following the start of ARVs [7]. The overall survival probabilities for the first and fifth years were 0.9376 and 0.8049, respectively. These findings, projected an improvement in terms of survival probabilities when compared with another study conducted in Ghana in the Lawra and Jirapa districts in the upper west region where the survival probability at the end of the three years was 0.795 per 1000 person years [7]. The findings were similar to the findings of a study conducted in the Henan province of China from 2005 to 2014 where the cumulated survival for the first and fifth year were 93.7% and 85.3% respectively [20]. The reason for the improvement in survival probabilities could be due to the removal of all barriers to the initiation of ARVs and early diagnosis due to the 90-90-90 policy which was not the case during the period of the study by Okyere et al., (2015).

Significant differences in survival probabilities were observed among specific variables using the Kaplan-Meier survival curve. WHO stages I and II had the highest survival probability and this finding agreed with studies in Ghana’s upper west region and Oromiyaa, Ethiopia where WHO stages I and II had the highest survival probability than stages III and IV [7,21]. The reason for the variation among the various stages could be due to the prognosis of the disease at the time of diagnosis and initiation of the ARVs. Although disclosure of one’s status may play a role in controlling the spread of HIV, it had very little impact on the improvement of the survival probability of the clients in the short term. However, in the long term, disclosing status disclosure gives an individual a better chance of survival compared to someone who has not disclosed the status. The reason for this dynamics could be due to the support that those who disclose their status receive from their treatment supporters. Females had better survival probabilities than their male counterparts and this can be attributed to the health-seeking behaviors of females [22].

The overall mortality rate observed was low compared with what was found in a retrospective cohort study from 2004-2013 in Nepal [23]. The reason for the reduction in overall mortality in this study may be due to the fact that majority of the participants were diagnosed at WHO stage I and II [24]. Mortality was highest among those diagnosed at the WHO stage III and IV, meaning this group will need special attention, especially within the first two years of diagnosis where most of the deaths occurred. Regular clinic visits, viral load monitoring, and adherence to the ARV regimen are essential to prevent mortalities. Other areas where significant variations in mortality was observed were gender and religion. Morality was high among males compared to females and that can be attributed to the health seeking behaviors of the male gender [22].

The hazard of mortality after the initiation of ARVs was statistically significant and associated with those in WHO stages III and IV and age group 50^+^ years and after adjusting in a multivariable analysis, WHO stage III and IV and age group 50^+^ years remained significant. The findings were similar to other studies that were conducted in Ghana, sub-Saharan Africa, and other parts of the world [7,17,21,23,25]. From the smooth hazard curve, the risk of mortality is highest in the first and second years of ARVs initiation. More pragmatic efforts including regular clinic visits, early diagnosis of opportunistic infections, viral load monitoring and so on, are required to reduce the death rate within the first and second years of ARVs initiation.

### Strengths and limitations

This is the first study of its kind on this subject and will contribute greatly to literature and also serve as a baseline finding for future studies. The lack of a comparison group restricted our ability to compare the outcome between those initiated on ARVs and those that are not on ARVs. Hence, comparison was done using previous studies while the presence of missing data also restricted the inclusion of variables such as viral load.

## Conclusion

Survival probabilities have improved significantly among PLWH in the two districts compared to previous studies conducted before the introduction of the treat-all policy. We recommend the need for early diagnosis through testing at all entry points to hospitals and clinics and through the “Know your Status” campaigns for HIV. Clients who are 50 years and above and those who were diagnosed at the WHO stages III andIV should be given shorter clinic visit intervals and ensure viral load monitoring is done as recommended by the National AIDS Control Program.

## Data Availability

All relevant data are within the manuscript and its Supporting Information files.

## Availability of data and materials

All data are fully available without restriction

## Competing interests

The authors declare that no competing interest existed for this study

## Funding

None

## Author Contributions

Conceptualization: ES, RNA, MMB, KB, MSY, DAO, AM, DB, EKN & AKE

Data curation: ES, RNA, NKAB & MMB,

Formal analysis: ES, RNA, MMB, KB & MSY

Methodology: ES, RNA, MMB, KB, MSY, DAO, NKAB, AM, DB, EKN & AKE

Project administration: ES, RNA, MMB, KB & MSY,

Supervision: AM, DB, EKN & AKE

Validation: DAO, AM, DB, EKN & AKE

Writing – original draft: ES, RNA, MMB, KB & MSY

Writing – review and editing: DAO, NKAB, AM, DB, EKN & AKE

## Acknowledgements

We are grateful to all PLHIV whose records we reviewed for this study. To all the staff of the four ARVs centers in both Tatale and Zabzugu, we thank them for their efforts in saving lives and the documentation without which this study would not have been possible. We are also grateful to the management of the two hospitals and the District Directors of Health Services for their approval of the study. We are most grateful to God for the inspiration, guidance and protection throughout the study.

## Notes

### Competing Interest Statement

The authors have declared no competing interest.

### Clinical Trial

"N/A"

### Funding Statement

The authors received no specific funding for this work.

